# Leveraging NLP to Identify Domain-Specific Variables in Large-Scale Cohort Metadata: A Sleep Use Case

**DOI:** 10.64898/2026.01.18.26344317

**Authors:** Ben Draper, Penelope Briggs, Shaun Purcell, Derk-Jan Dijk, Sarah Bauermeister, Ullrich Bartsch

**Affiliations:** Department of Chemical, Materials & Biological Engineering, University of Sheffield; Surrey Sleep Research Centre, University of Surrey; Department of Psychiatry, Brigham & Women’s Hospital, Harvard Medical School; UK DRI Care Research & Technology Centre; Department of Psychiatry, University of Oxford

## Abstract

Public health policies increasingly rely on the use of complex and large datasets containing heterogeneous, multimodal data that require advanced analytical methods to extract meaningful insights and support evidence-based decision-making. Essential for the sharing and analysis of public health data is the description of the data (“data about data”) or metadata. Indeed, a lack of metadata standards has been identified as a key technical barrier to public health data sharing ^1^.

Metadata varies considerably between cohort studies. It often contains extremely heterogenous variable and data descriptions for similar or identical metrics. The assessment and processing of metadata is very labour intensive. It relies on researchers manually sifting through large numbers of variable descriptors and other study data documentation to extract variables of interest. Unless a validated quantitative tool is used, the variability in phrasing is surprisingly high. Variations in metadata can makes it hard to find and compare results across studies.

Specifically, questions about wake-sleep behaviour like subjective sleep quality and duration are commonly employed in large cohort studies – but vary not only in the wording of the questions put to participants but also in the metadata that describes the questions and their responses.

We developed a semi-automatic retrieval method (METAMATCH) for sleep-related variables from metadata obtained from multiple large cohort studies. Here we employ sleep as the health area of interest, but in principle this method can be applied to any area of research. We developed the retrieval method using metadata provided by Dementias Platform UK (DPUK, https://www.dementiasplatform.uk/) a Trusted Research Environment (TRE) that hosts over 50 cohort datasets of different sizes. From this we extracted a metadata corpus describing 86,682 variables across 17 cohort studies.

To identify sleep-related variables, we curated a reference dictionary (the SLEEPTALKING corpus) combining terms from the Unified Medical Language System (UMLS) and expert knowledge. We then employed Term Frequency-Inverse Document Frequency (TF-IDF) vectorisation and cosine-similarity to rank variable-descriptions by comparing the metadata corpus to the SLEEPTALKING corpus. We identified 337 semantically unique sleep-related variables. We then performed semantic analyses to characterize these variables. We applied Bidirectional Encoder Representations from Transformers (BERT) embeddings and unsupervised cluster analysis. The results indicate that definitions tend to group by cohort and length of the variable description. We validated the cluster analysis using expert-based categorisation of variable descriptions. The most common categories identified were ’Sleep Difficulty,’ ’Sleep Duration,’ and ’Sleep Latency & Waking.’

Consequently, expert knowledge and consensus remains essential for accurately categorizing variables within a universal ontology across different studies. We conclude that combining NLP techniques with expert input offers an efficient approach to harvesting metadata across multiple cohort studies.

## Introduction

Sleep is a universal behavioural state of rest during which consciousness is suspended, and interaction with the environment is reduced. Sleep and overall well-being are closely tied, with good sleep hygiene being tied to better mental and physical health. Abnormal sleep behaviour can serve as an important indicator of disease risk or as a symptom of an existing condition. Among diseases, neuropsychiatric disorders are notably influenced by sleep in a bidirectional relationship, where symptoms and risks overlap.

Validated quantitative instruments for clinical sleep research include the Pittsburgh Sleep Quality Index (PSQI), which assesses sleep quality over a month, including duration, latency, and disturbances ^2^. The Epworth Sleepiness Scale (ESS) measures daytime sleepiness, often linked to conditions like sleep apnea ^3,4^, while the Insomnia Severity Index (ISI) evaluates the impact of insomnia on daily life ^5^. Other questionnaires, such as the Berlin Questionnaire for obstructive sleep apnea (OSA), the Sleep Hygiene Index (SHI) for sleep habits, and the Holland Sleep Disorders Questionnaire (HSDQ) for a broad range of sleep disorders, are also widely used ^6–8^. In addition, instruments that support psychiatric diagnoses often include sleep-related questions, either as standalone items or within sections on overall health. For instance, the Beck Depression Inventory (BDI), a tool for assessing depressive symptoms, contains several questions related to sleep disturbances ^9^. Similarly, general health questionnaires like the General Health Questionnaire (GHQ) and Short Form 36 (SF-36) include sleep-related questions ^10,11^. In large population studies, these questionnaires have proven useful in identifying links between sleep and neuropsychiatric disorders^12,13^.

But many current and historical studies often rely on non-standardised approaches to collect information on sleep. In the UK, several large cohort studies have collected population-wide health insights over time, for example the English Longitudinal Study of Ageing (ELSA), Generation Scotland, and the Cognitive Function and Ageing Studies (CFAS) and more recently the UK Biobank. These studies have spanned multiple decades, gathering valuable data on health and disease across the lifespan^14–16^. Population sizes in these studies vary widely, with the UK Biobank involving over 500,000 participants, ELSA starting with over 11,000 participants, Generation Scotland including over 24,000, and CFAS I and II covering multiple waves of data collection from tens of thousands of individuals. But the amount of information on sleep is not easily identifiable in these cohorts.

In principle the availability of multiple large cohort datasets inside a TRE lends itself to pooling of individual-level data^17^ as an alternative to the more common meta-analysis of exposure-outcome associations across studies. One significant challenge in conducting pooled analysis is the labour-intensive process of data curation and harmonisation, which is hindered by high variability in the associated metadata.

Natural Language Processing (NLP) offers a potential solution to some of these challenges by extracting meaningful information from unstructured data. Key components of NLP include information extraction, text classification, and semantic similarity assessment. It has been successfully employed for the processing of Electronic Health Records (EHRs)^18^. But NLP is rarely used to efficiently process and categorise metadata in large cohort studies.

In this study, we applied a text-mining approach to align sleep-related data from DPUK cohorts with standardized terminologies from the Unified Medical Language System (UMLS). To enhance classification, we employed unsupervised clustering on transformed word embeddings, enabling a robust, data-driven exploration of sleep-related definitions across the cohorts. This methodology facilitated comprehensive harmonization of the data, bridging diverse representations of sleep variables for prospective analysis.

## Methods

We extracted sleep-associated variables from cohort study metadata using a curated reference dictionary (the SLEEPTALKING corpus). We computed variable definition similarity, ranked the scores and performed manual quality control.

### Selected cohort study metadata

We used a subset of cohorts available at the DPUK Data Portal to identify sleep variables. DPUK has an existing ontology for metadata^19^, but sleep questions are often dispersed across multiple categories of related data, such as physical, psychological or mental health questionnaires.

20 cohorts were selected and approved for a detailed analysis of their associated metadata. We included AMPLE, Airwave, BDR, BRACE, CamCAN, CamPaIGN, CaPS, CFAS I & II, ELSA, EPIC, EPINEF, GS: SFHS, ICICLE, MRC, NICOLA, NIMROD, SMC, TRACK and Whitehall respectively ^20–38^. These are a mixture of cross-sectional and longitudinal studies. Some of the longitudinal studies are continuously updated and subject to new waves of data. All cohort full names, dates, summary description and references are listed in **Supplementary Table 1**.

### Reference dictionary – SLEEPTALKING corpus

The Unified Medical Language System® (UMLS®), developed by the National Library of Medicine (NLM), is a comprehensive framework that standardizes biomedical terminologies, classifications, and coding systems to enhance integration and interoperability. In this study, we employed the UMLS Metathesaurus, which links synonymous biomedical terms from over 200 source vocabularies, to extract sleep-related concepts via the UMLS API. Medical vocabularies such as ICD-10 and SNOMED CT were unified under common concept unique identifiers (CUIs). Using the parent CUI C0037313 for “Sleep,” 2032 sleep-related definitions were retrieved from the UMLS Metathesaurus in June 2022. The top five vocabularies where sleep-related definitions were found include LOINC®, MEDCIN®, SNOMED CT®, UMLS Metathesaurus, and the NCI Thesaurus.

### Identifying variables of interest

Metadata from 20 cohorts were manually converted from original file formats (XML, XLSX or DTA) and collated into a single CSV file. The variable descriptors were transformed into N-grams - sequences of consecutive word tokens or symbols that capture patterns, including similar word roots. To enhance relevance, inconsequential words (known as STOP words) were removed. The Term Frequency-Inverse Document Frequency (TF-IDF) method was then applied to create N-gram-based vectors, measuring the importance of each word relative to the entire metadata set.

We then calculated the cosine similarity for each vector pair. The cosine similarity quantifies the distance between two vectors based on the cosine of the angle between them, with scores ranging from −1 (completely different) to 1 (completely identical), and 0 representing no similarity.

The cosine similarity is defined as

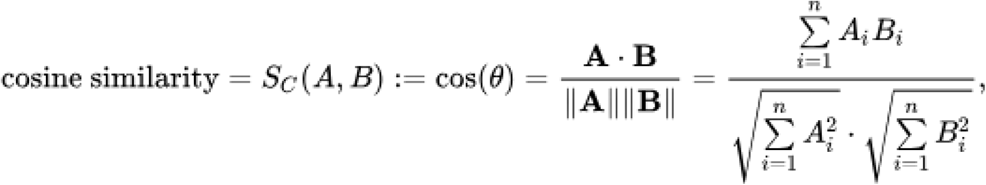

• 𝐴 ⋅ 𝐵 represents the dot product between the two vectors.

• ∥ *A* ∥ *or* ∥ *B* ∥ is the Euclidean norm of vector A or B.

We set

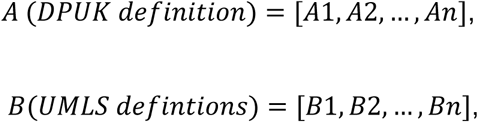

Cosine similarity scores of each vector pair were used to map each definition to the best-matching UMLS term.

These scores ranged from 1.0 (perfect match) to 0.164 (low similarity), with an average of 0.522. The scores were then manually reviewed to verify whether sleep-related variables had been properly identified. Initially, we identified 754 sleep-related definitions from 86,682 variable descriptions.

### Semantic analysis & refinement using natural language processing (NLP) techniques

All variable descriptors (nv_total_=86,682) were converted into word embeddings to evaluate semantic similarities between sleep and other variables and to explore linguistic diversity across cohorts. From 754 sleep-related variables, 337 unique definitions were distilled for further analysis. Dense vector representations capturing the semantic meaning of each definition were generated using Bidirectional Encoder Representations from Transformers (BERT) embeddings, specifically MiniLM-L6-v2 from HuggingFace, with the CLS “classification” token summarizing sentence meaning. BERT embeddings are contextualised word representations generated by the BERT model, capturing the meaning of words based on their surrounding context in a sentence ^39^. These embeddings were then visualised using t-distributed Stochastic Neighbour Embedding (t-SNE), to identify clusters and explore the local grouping behaviour of the tokens in two dimensions.

A manual retrieval of full sentence definitions from supplementary metadata provided additional definitions which were added to the retrieved list. This final list of definitions was explored further. For explorative clustering, the HDBSCAN (Hierarchical Density-Based Spatial Clustering of Applications with Noise) method was used to help visualise groups ^40^. HDBSCAN is particularly suitable for clustering BERT CLS embeddings in linguistic tasks because it can effectively handle high-dimensional complex structures, which often have varied cluster densities due to the diverse meanings and nuances of language. Its ability to discover clusters without a predefined number and to manage noise is optimal when working with linguistic data which typically contains both well-defined concepts and ambiguous, less-represented instances. This adaptability makes HDBSCAN ideal for clustering semantic representations derived from BERT’s contextualised embeddings.

### Manual categorisation of definitions

A manual categorisation process was employed to group and label the sleep definitions into relevant categories for prospective researchers. We assigned 15 main categories which consisted of 14 cluster-based categories and one validated instrument category (**See Table 2**). Validated instruments included 1) The Pittsburgh Sleep Quality Index (PSQI): a self-report questionnaire used to assess sleep quality and disturbances over 1 month 2) The Epworth Sleepiness Scale: a self-assessment tool designed to understand how daytime sleepiness impacts daytime function. It consists of eight questions; each answered on a four-point scale. 3) The Center for Epidemiologic Studies Depression Scale (CES-D): a tool used to measure depressive symptoms which includes questions on sleep. 4) The General Health Questionnaire GHQ: a screening tool designed to detect psychological distress and potential psychiatric disorders in various settings.

Using the keywords exemplified in **Table 2**, the definitions were categorised and verified by each of the main authors through manual consensus.

### Software & Packages

All analyses were conducted using R (version 4.4.1) and custom Python (version 3.14) scripts. The majority of the analysis was performed using R, with a single Python script employed to generate BERT embeddings from the metadata. Below is a description of the key packages used in Natural Language Processing (NLP) and clustering, with a comprehensive list available in the Supplementary Information.

1. **BERT Embeddings** To convert metadata definitions into vector representations, we used the ‘transformers’ Python package (version 4.44.0). Specifically, the BertTokenizer and BertModel classes were utilized to tokenize and compute the BERT-based embeddings for the metadata definitions. These embeddings capture rich contextual information, essential for subsequent analysis.
2. **Cosine Similarity Calculation** For calculating cosine similarity between the extracted DPUK metadata definitions and UMLS definitions, we used the R package ‘reticulate’ (version 1.4.0). This package enables seamless interaction between R and Python, allowing us to utilize the ‘polyfuzz’ (version 0.4.2) Python package. Polyfuzz employs TF-IDF-based k-nearest neighbors (k-NN) matching to compute cosine similarity scores. This process matches similar definitions between the two datasets based on their textual content.
3. **Dimensionality Reduction and Clustering**

a. Non-linear Dimensionality Reduction For the initial dimensionality reduction of the extracted word embeddings (which represent sleep definitions), we employed the R package ‘Rtsne’ (version 0.17). This package first applies Principal Component Analysis (PCA) to reduce the data to a lower-dimensional space, followed by t-SNE (t-Distributed Stochastic Neighbour Embedding) to further reduce the dimensionality to 2D. The parameters used for t-SNE were: dimensions = 2 and perplexity = 30. t-SNE was used to visually identify potential similarities between the word embeddings of the semantically unique sleep definitions.
b. Uniform Manifold Approximation and Projection (UMAP) Following t-SNE, we applied further dimensionality reduction technique, UMAP (version 0.2.10.0), using the R package ‘umap’. We set the parameters for UMAP as follows: n_neighbors = 10, min_dist = 0.01, n_components = 5, and used the cosine metric to measure distances between points. UMAP is known for producing more interpretable embeddings (better preserving global structures), particularly when the data contains complex relationships such as in linguistic semantics.
4. **Unsupervised Clustering with HDBSCAN** To perform unsupervised clustering on the word embeddings, we utilized HDBSCAN (Hierarchical Density-Based Spatial Clustering of Applications with Noise) implemented in the ‘dbscan’ R package (version 1.2.0). HDBSCAN was applied to the UMAP embeddings to group the semantically unique sleep definitions into five distinct clusters. HDBSCAN is particularly suited for linguistic data, as it excels at clustering high-dimensional data while handling noise effectively. It works by identifying dense regions in the data and separating them from sparser areas, thus preserving the hierarchical structure of clusters.

**A full package list is included in the supplementary information.**

## Results

### Cohort studies are diverse in population and variable size

Prior to metadata mining, we performed a descriptive analysis of cohort variable definitions. We analysed metadata from 20 cohorts with estimated total participants of 178,978 (N_total_). Cohorts showed significant variation in participant numbers, ranging from as few as 80 participants in the AMPLE cohort to as many as 53,280 in the Airwave cohort.

The metadata comprises 86,682 variables (nv_total_), with the largest proportion (52%, nv = 44,826) derived from the longitudinal ELSA dataset. In contrast, the single wave CamPaIGN cohort contributes less than 1% (nv = 32) of the variables. Together, the top three cohorts ELSA, CFAS, and CaPS represent 91% of the variables (nv = 78,275) in the metadata, yet they represent only 18% (N = 33,063) of the number of participants across all cohorts (**see Figure 1 a and b**). Three cohorts, Airwave (N = 53,280), GS-SFHS (N = 23,960), and EPIC (N = 25,639) together account for approximately 57% of all participants but comprise only 2% of the number of variables in the metadata (nv = 1745). This fact highlights that the number of participants in a cohort does not directly predict the number of variables in a study.

**Figure 1.**
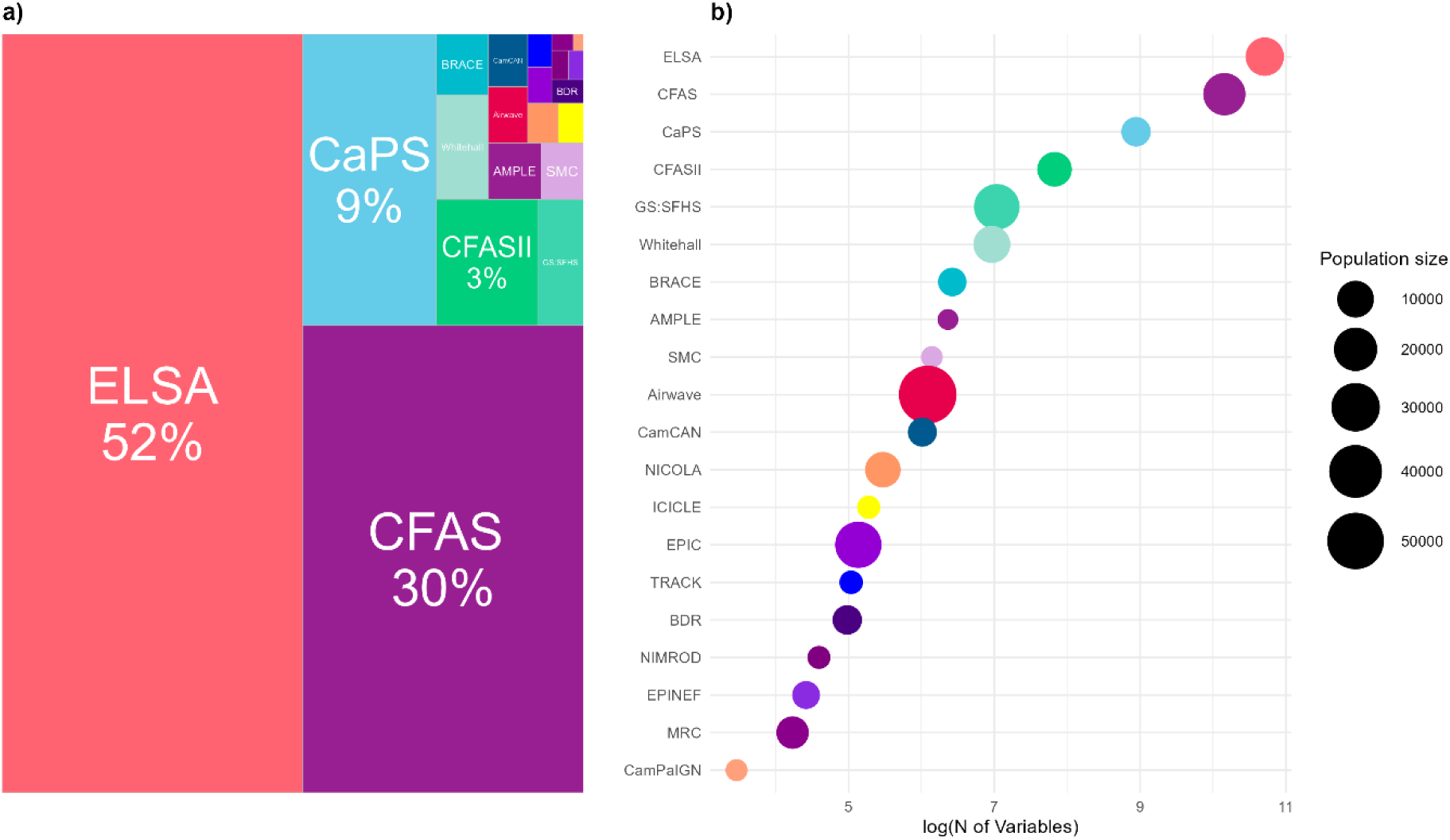
a: Distribution of cohort variables within the collected meta-dataset. A treemap of the meta-dataset comprising 86,682 variables. Area of square represents the proportional number of variables of the entire dataset. **b: Relationship between cohort size and number of variables.** The x-axis shows the logarithm of the number of variables per cohort, with bubble size indicating the total number of participants. The plot demonstrates that cohort size does not directly correlate with the number of variables, as seen in the varying bubble sizes across different positions on the x-axis.

### Sleep variable identification

To identify sleep-related variables from the cohort metadata, we employed the TF-IDF cosine similarity to compare metadata definitions with the SLEEPTALKING corpus. We identified 754 variables with similarity scores ranging from 1.0 (perfect match) to 0.18, which for simplicity we will refer to as ‘sleep variables’. Of the 20 original cohorts, 3 (EPINEF, BDR and TRACK) did not contain any sleep variables. After an initial identification of sleep related variables, we removed duplicates (re-used variable descriptions across multiple waves within a study) (**see Figure 2 a**). An example duplicate was the CFAS description “Any trouble sleeping?” as variable v113_x which has 11 instances across waves (where x = c12, c14, c16, c2, c6, c8, cx, f1, f3, s6, w8). Duplicates were most found in the CFAS I, CFAS II and CaPS cohorts, all longitudinal studies with data collected over multiple time points (CFAS I:1989-2011, CFAS II:2008-ongoing, CaPS:1979-ongoing) (**see Figure 2 b**). For example, ICICLE has over 50% duplication, in a small absolute number of variables. Duplication is minimal in ELSA and MRC while the remaining 11 cohorts have no duplicates.

**Figure 2.**
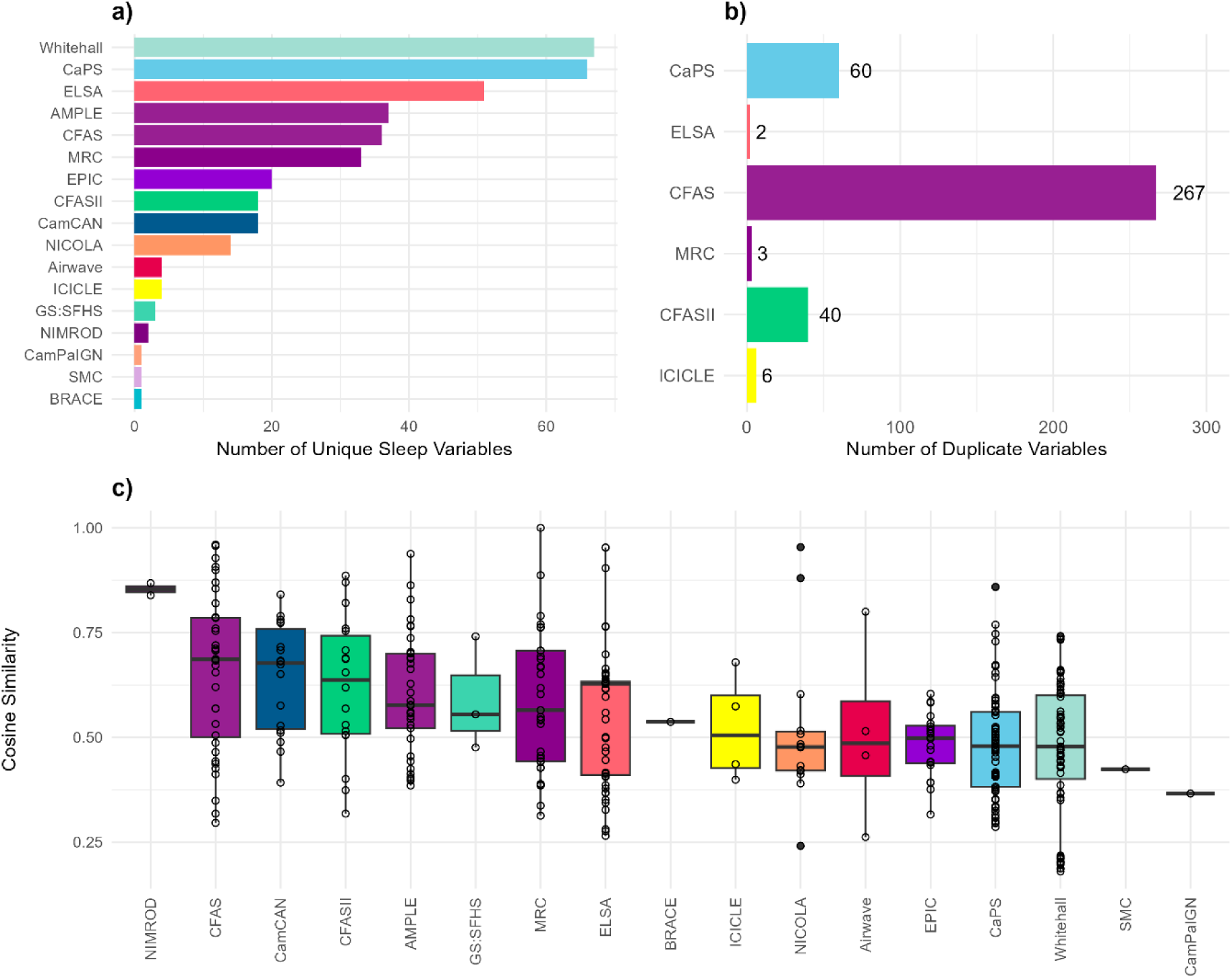
**a: Number of unique sleep variables across different cohorts**. This bar chart shows the number of unique variables (n=376), defined by one identifier for one exact definition. **b: Number of duplicated sleep variables across different cohorts**. This bar chart shows the number of duplicated variables (n=378), defined by multiple identifiers for the same exact definition. Although duplication is minimal in most cohorts, it is notably higher in long-term longitudinal studies such as CFAS, CFASII, and CaPS, where repeated data collection over time has led to multiple identifiers for the same variables. **c: Distribution of similarity scores to UMLS sleep definitions across 17 remaining cohorts ranked by their average score.** The scores vary widely, with NIMROD achieving the highest average similarity at 0.868 (very high), while CamPaIGN possesses the lowest at 0.374 (medium low). Each box represents the interquartile range (IQR) of cosine similarity scores, with the central line indicating the median. Outliers and variations within the cohorts highlight differing degrees of alignment between cohort-specific sleep definitions and the UMLS standard.

After removing duplicates, we continued to process 376 unique sleep descriptions across the remaining 17 cohorts for further semantic analysis. These unique variable definitions were scored based on their TF-IDF cosine similarity to UMLS definitions and ranked by their average score (**see Figure 2 c**). While the maximum possible score was 1.0, the highest cohort average score was 0.868, achieved by NIMROD with its two variable matches and the lowest for CamPaIGN at 0.374.

To illustrate the range of similarities of variable definitions compared to the SLEEPTALKING corpus, we describe examples of high and low-scoring definitions (**Table S2**). The highest-scoring definitions were typically short and semantically aligned closely with the UMLS wording. For example, the CFAS definition “s. has difficulty in sleeping?” scored 0.96, which was nearly identical to the UMLS definition “difficulty sleeping”. This high similarity demonstrated how succinct, clear, and directly comparable definitions are almost identical to UMLS wording. In contrast, lower-scoring definitions, like Whitehall’s “q2.64d wake as usual but tired (s5),” which scored only 0.18, contained additional contextual or numerical information that reduced their semantic alignment with the UMLS wording. Additional examples of high and low-scoring definitions are shown in **Table S2**. For instance, the Whitehall variable MSLEEP, defined as “q72 week-night average hours of sleep (s7),” received a TF-IDF score of 0.454 when matched to the UMLS term “Shortened sleep cycle (average 5.7 hours per night),” highlighting the difficulties in accurately aligning more complex variable definitions with UMLS terms. This pattern suggests that simpler, less complex variable definitions tend to produce more accurate matches with the SLEEPTALKING corpus.

This initial survey demonstrates the value of the SLEEPTALKING corpus for identifying sleep-related variables and its ability to accelerate harmonization efforts across heterogeneous datasets and advancing epidemiological sleep research.

Our approach highlights the potential of UMLS terms for categorizing metadata, especially when variable definitions are concise and closely aligned with standardized wording. For cohorts with more complex or detailed definitions, additional refinement or preprocessing steps could further enhance alignment with UMLS terms, opening new approaches to aid harmonization across diverse datasets.

### Semantic Analysis of Sleep Variable Descriptions

Next, we investigated the semantic relationships between the variable descriptions. We conducted a semantic analysis using the pretrained ’bert-base-uncased’ model, we processed the original metadata set of 86,682 variables to generate BERT word embeddings. After text preprocessing (e.g., removing punctuation, parentheses, and redundant patterns), we extracted the [CLS] token embedding for each cleaned definition, yielding 35,529 unique embeddings representing semantically distinct variable definitions across the DPUK cohorts which were utilised for subsequent analysis.

To capture the semantic landscape, we reduced the 35,529 unique word embeddings, using the [CLS] token as a summary representation of each definition. To evaluate whether sleep-related variables exhibited distinct patterns relative to the broader metadata, we applied t-Distributed Stochastic Neighbor Embedding (t-SNE) to visualize CLS token similarity (**Figure 3a**). In the t-SNE plot, sleep variables (red dots) were dispersed across the plot without forming clear clusters, indicating that BERT embeddings captured semantic relationships but did not strongly differentiate sleep-related variables from others. This suggests significant semantic overlap between sleep and non-sleep metadata in the high-dimensional space. Focusing on just the “semantically unique” sleep variables (n = 337 / 376), we performed a second t-SNE analysis on their CLS tokens (**Figure 3b**). This revealed clustering of sleep-related definitions by cohort, driven primarily by the length and complexity of the definitions rather than their content. Cohorts with similar linguistic styles formed distinct groups, highlighting that cohort-specific language significantly influences the semantic organization of sleep variables.

**Figure 3:**
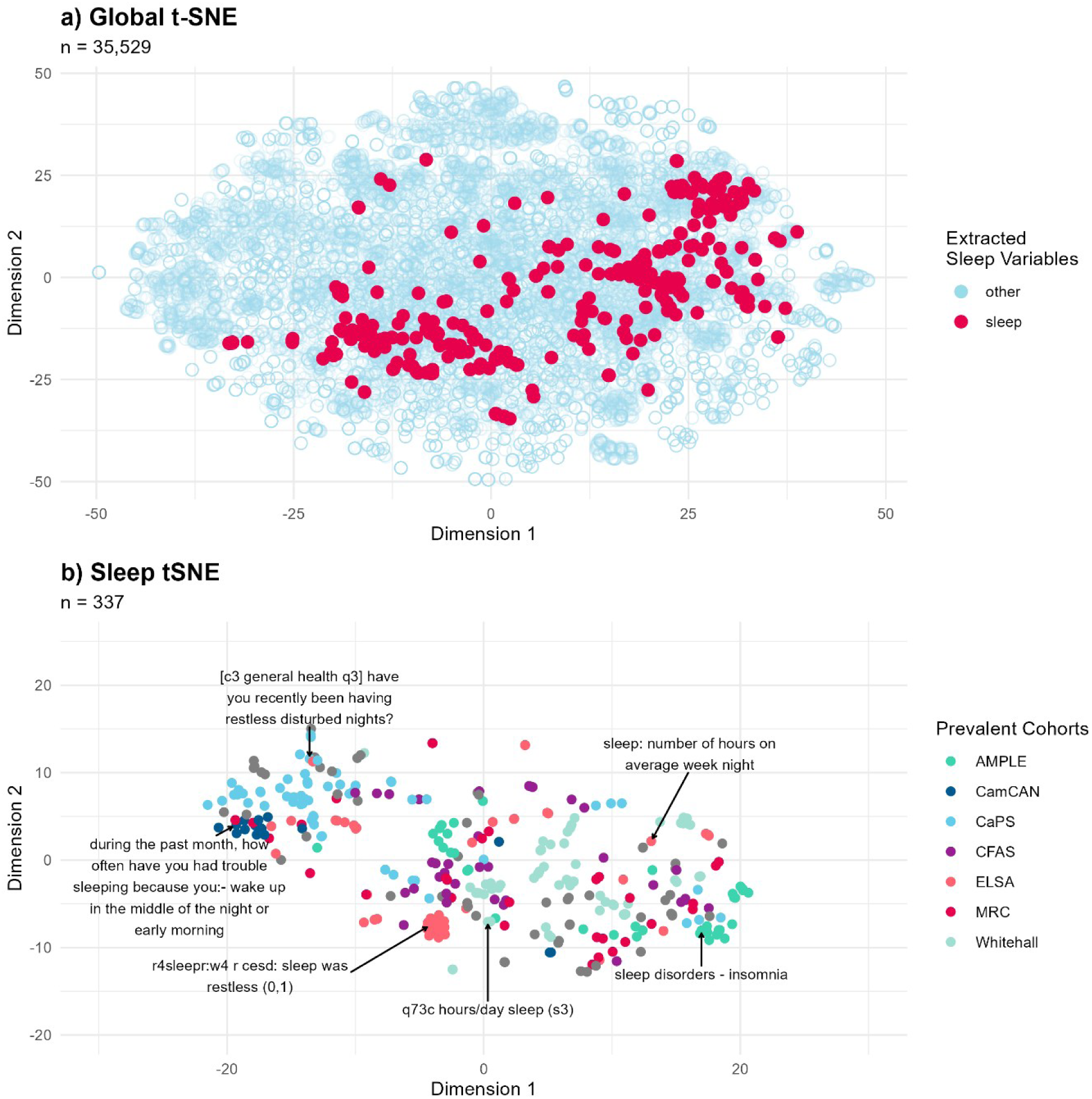
a: t-SNE visualization of BERT CLS embeddings for 35,529 unique cohort definitions. The plot shows the clustering of sleep-related variables (red) as distinct from other variables (light blue), indicating semantic grouping of sleep variables within the metadata. **b: t-SNE visualization of BERT CLS embeddings for 337 semantically unique sleep-related cohort definitions.** The plot showed clustering based on cohort-specific language and the complexity of sleep definitions.

Cohorts using similar language formed distinct groups, indicating that linguistic differences, rather than the data content itself, drove the observed clustering in the semantic analysis. This suggests that cohort-specific language plays a significant role in how sleep variables are interpreted and organized within the semantic space.

### Metadata may be insufficient

The t-SNE visualization underscored a key distinction between longer, questionnaire-style definitions and shorter, semantic questions, which accounted for the primary variation in the BERT embeddings. To address this, we manually reviewed and expanded the metadata utilising the full-format questionnaire questions sourced (where available) from the DPUK portal and cohort websites. This step was critical, as the original metadata often lacked sufficient context. For instance, the CFAS definition “s. wake early in morning?” was expanded to its full question: “Does s/he wake early in the morning, before his/her normal time, and not get back to sleep again?”. This expansion, notably in CFAS studies, clarified semantic intent not evident in the abbreviated metadata. Overall, we expanded only 19% (73/376) of the original extracted sleep metadata descriptions to include the full format questions. However, documentation remained limited for some cohorts (e.g., AMPLE, SMC, Whitehall), constraining full resolution.

Using these updated full definitions, we reran BERT embeddings, reduced them to four dimensions with Uniform Manifold Approximation and Projection (UMAP), and clustered them using HDBSCAN, yielding six meaningful clusters (**See Table 1 and Figure 4a**). These clusters approximated common category labels but were imprecise, with numerous outliers, reflecting the challenges of clustering linguistic data for precise grouping. While unsupervised clustering of BERT embeddings provides thematic insights, our findings indicate it falls short of capturing true semantic meaning without expert consensus. This underscores that metadata alone may be insufficient for accurate categorization of sleep-related data, necessitating contextual expansion to full-format questions for robust analysis.

**Figure 4:**
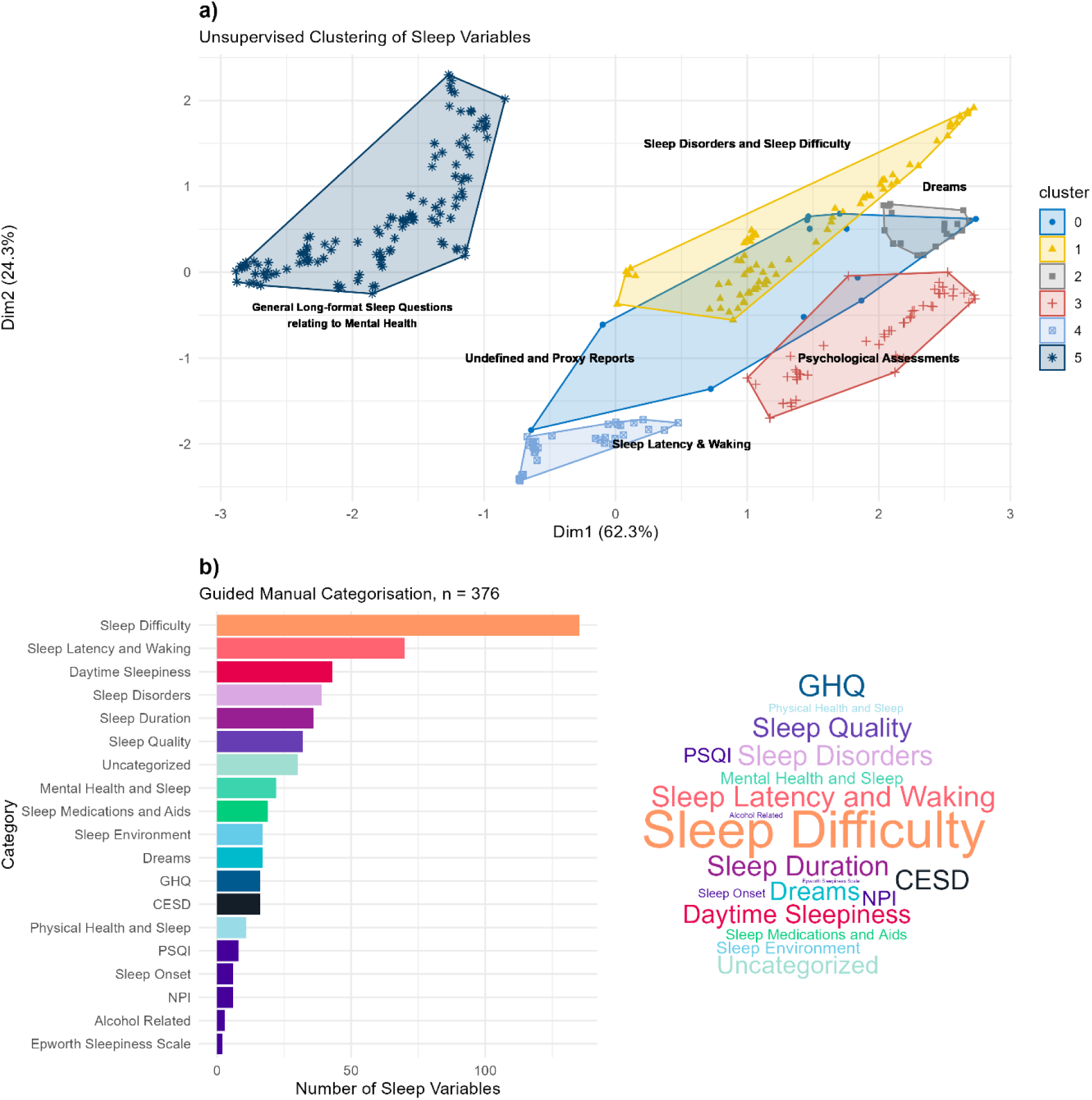
a: HDBSCAN Clustering of UMAP-Reduced BERT Embeddings. This plot shows the thematic clustering of BERT embeddings using UMAP and HDBSCAN. Clusters represent manually verified categories such as General Sleep Patterns, Sleep Disorders, Daytime Sleepiness, Sleep Latency, Chronic Sleep Difficulty, Psychological Assessments, and the impact of Mental Health on Sleep. Cluster boundaries are represented by ellipses, with shapes and colours corresponding to different clusters. **b: Sleep Categories extracted from the metadata**. The bar chart shows the number of unique sleep variables within categories ordered by count (x axis).The word cloud visualizes the distribution of categorized sleep-related variables, with the size of each category’s name reflecting the frequency of variables within each category. […]

**Table 1:** Data Driven Linguistic Clusters of Sleep Definitions. Results from the unsupervised HDBSCAN clustering of BERT embeddings reveals 6 distinct clusters with an uncategorised separate cluster.

| Cluster # | Cluster description | Comments |
| --- | --- | --- |
| 0 | Uncategorised general sleep-related data and proxy-reported behaviours | Often derived from indirect observations |
| 1 | Sleep disorders such as Nocturia, sleep apnoea, insomnia, and restless leg syndrome, | Highlights difficulties falling asleep and related disorders |
| 2 | Dreams | Reflects dreams, nightmares, parasomnia or sleep paralysis. |
| 3 | Shorter psychiatric assessment definitions | Includes scoring from clinical tools such as NPI, CES-D, and GHQ. |
| 4 | Undefined & Proxy Reports | Uncategorised or proxy reporting where someone is describing some else's sleep patterns. |
| 5 | Longer definitions focused on the interaction between mental health and sleep | Describes the impact of mental health conditions on sleep quality and duration. |

**Table 2:**
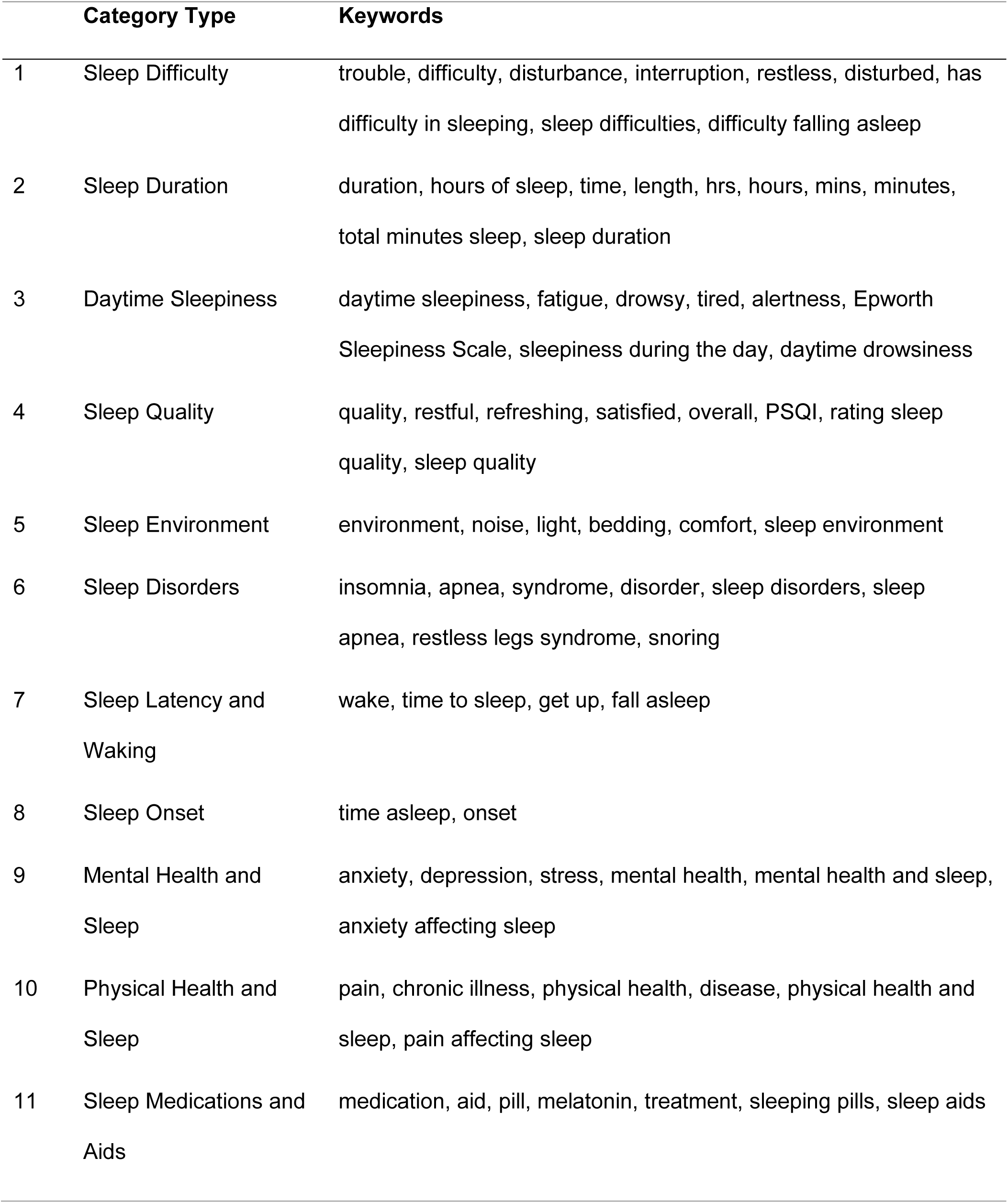
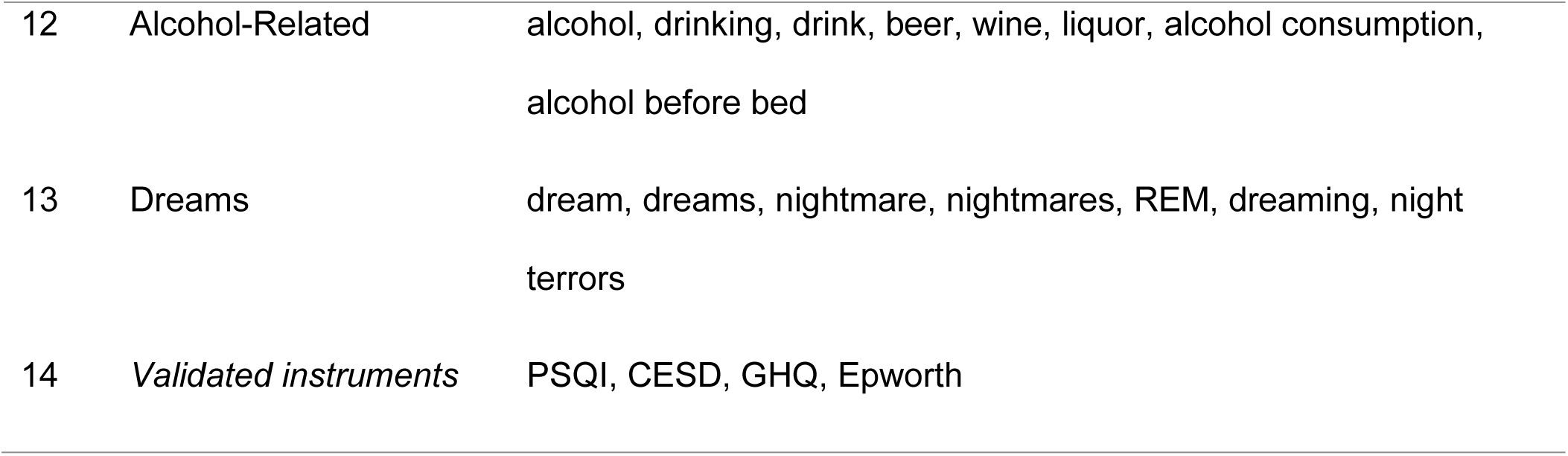
Keyword-Based Categories for Sleep-Related Variables. The table lists the categories used to categorise sleep-related variables, along with associated keywords. These keywords were used to scan the dataset and automatically assign variables to relevant categories which were then manually curated on consensus.

|  | Category Type | Keywords |
| --- | --- | --- |
| 1 | Sleep Difficulty | trouble, difficulty, disturbance, interruption, restless, disturbed, has difficulty in sleeping, sleep difficulties, difficulty falling asleep |
| 2 | Sleep Duration | duration, hours of sleep, time, length, hrs, hours, mins, minutes, total minutes sleep, sleep duration |
| 3 | Daytime Sleepiness | daytime sleepiness, fatigue, drowsy, tired, alertness, Epworth Sleepiness Scale, sleepiness during the day, daytime drowsiness |
| 4 | Sleep Quality | quality, restful, refreshing, satisfied, overall, PSQI, rating sleep quality, sleep quality |
| 5 | Sleep Environment | environment, noise, light, bedding, comfort, sleep environment |
| 6 | Sleep Disorders | insomnia, apnea, syndrome, disorder, sleep disorders, sleep apnea, restless legs syndrome, snoring |
| 7 | Sleep Latency and Waking | wake, time to sleep, get up, fall asleep |
| 8 | Sleep Onset | time asleep, onset |
| 9 | Mental Health and Sleep | anxiety, depression, stress, mental health, mental health and sleep, anxiety affecting sleep |
| 10 | Physical Health and Sleep | pain, chronic illness, physical health, disease, physical health and sleep, pain affecting sleep |
| 11 | Sleep Medications and Aids | medication, aid, pill, melatonin, treatment, sleeping pills, sleep aids |
| 12 | Alcohol-Related | alcohol, drinking, drink, beer, wine, liquor, alcohol consumption,<br>alcohol before bed |
| 13 | Dreams | dream, dreams, nightmare, nightmares, REM, dreaming, night<br>terrors |
| 14 | *Validated instruments* | PSQI, CESD, GHQ, Epworth |

### Categorisation of sleep-related variables

We categorized the 376 sleep definitions into groups based on keywords to determine the distribution of variables across different themes. This categorisation procedure aimed to organize a dataset of sleep-related variables from various cohorts using a systematic keyword-based approach, and 15 categories were identified.

However, 33 variables could not be categorized due to the absence of probable matches or insufficient information. An example metadata item is from CaPs: “[c3 sleep disturbance] the unique number allocated to each subject in the study as recorded on card 9”.

Keywords associated with these categories were manually scanned in the dataset fields, automatically grouping similar variables under relevant themes (**Table 2**). This approach also ensured the accurate categorisation of entries related to standardized sleep assessment tools, resulting in a more organized and interpretable dataset. The keyword-driven categorisation enhanced the dataset’s clarity and utility, making it more suitable for further analysis.

A word cloud **(Figure 4b)** was prepared of the 18 prominent sleep variables plus the uncategorised category, with the size of each category’s name reflecting the frequency of variables within each category (**see Table S3**). It distinguishes between generic categories (broadly defined themes) and specific categories (standardized assessment tools). As expected, the most prominent category relates to the broad concept of general “sleep difficulty”. This is because much of the sleep-related data involves respondents reporting varying levels of disturbance, interruption, or vague difficulties with sleep, without delving into specific issues. Categories like “Sleep Latency and Waking” and “Sleep Duration” may be more useful, as they represent non-subjective, quantifiable variables. Meanwhile, categories focused on specific sleep disorders, such as sleep apnoea, insomnia, or restless leg syndrome, provide more detailed insights into the underlying sleep issues.

## Discussion

In this study, we used metadata from 17 cohort studies available through DPUK to identify 337 semantically unique definitions of sleep-related variables.

Our approach employed standardized medical terminology ontologies available through UMLS including SNOMED CT and ICD, to create the SLEEPTALKING corpus and used NLP techniques to assess similarity between the SLEEPTALKING corpus and extracted variable definitions.

Semantic cluster analysis showed sleep variable definitions were clustered primarily by cohort, which suggests higher similarity of variable-definition style rather than variable-definition meaning, i.e. the metadata terminology ‘style’ used in a specific cohort study strongly influences the clustering of metadata word embeddings. Downstream manual ontological labelling of sleep variable definitions revealed “Sleep Difficulty” was the most prevalent category, followed by “Sleep Latency and Waking” and “Sleep Duration”. Our work demonstrates the feasibility of using existing ontologies to map metadata collections and identify variables of interest.

Our study is related to previous efforts for harmonising and annotating metadata, some of which included NLP or AI based tools^41^.

NLP frameworks have been adopted to extract clinical insights from complex cohort studies datasets and electronic health records (EHRs) collections ^42^. These included simplification of diverse tasks such as clinical named entity recognition, medical text classification, translation, and information retrieval ^43^.

Mc Elroy et al. employed word embeddings, semantic similarity, and rule-based curation to successfully pool mental health questionnaire data, mirroring aspects of our methodology^44^.

Dylag et al. employed pretrained large language models (e.g., BERT) to automatically semantically cluster variable definitions from the ELSA cohort. They achieved modest success in generating homogenous and complete clusters of metadata classes, underscoring persistent challenges in achieving robust clustering without human-assisted curation ^45^.

In addition, large language models (LLM) have recently been employed for harmonizing biomedical text and/or metadata ^43,45,46^. For example, GPT-4 achieved high accuracy in generating semantic labels of a thematic collection of patent texts, closely aligning with human annotations as measured by BLEU scores (Bilingual Evaluation Understudy) ^47^. This suggests that LLMs could address limitations of pure NLP approaches, by producing richer and more consistent metadata, especially when combined with full access to all additional study documentation including full questionnaires and study protocols.

In our view, enhancing the process of sleep related metadata extraction and classification, should include the following considerations.

First, a deeper linguistic understanding of sleep related definitions is needed to extract their semantic meaning and relationships. This could be achieved by fine-tuning LLMs. Mohseni and colleagues have outlined a systematic review of transformer architectures used in medical data analysis and highlighted several studies that have fine-tuned architectures such as BERT for medical record processing tasks ^48^. For instance, ClinicalBERT is specifically fine-tuned on clinical notes from the MIMIC-III database and has shown improved performance in predicting hospital readmissions and other clinical tasks compared to standard BERT ^49^. BioBERT is a pre-trained biomedical language representation model developed for biomedical text mining, utilizing a domain-specific corpus that includes PubMed abstracts and full-text articles ^50^. Utilising a domain-specific trained LLM may improve the clustering of sleep-related definitions and improve categorisation efforts.

Second, greater emphasis on expanding rule-based systems may help to improve the precision of information extraction. For example, a rule-based Named Entity Recognition (NER) approach can reliably extract relevant information from clinical documents, enhancing accuracy and efficiency. ^51^.

### Outlook

In summary, we created a manually curated set of sleep-related variables present in 17 longitudinal and cross-sectional studies that can be used by researchers to explore the relationship between sleep and other health outcomes. As other cohort studies continue to be added to the DPUK data portal the sleep related curation set can easily be expanded through repeated metadata analysis.

Future work will include the development of Shiny App for the R environment to enable semi-automated curation process inside the DPUK data portal. We envisage that an expanded SLEEPTALKING corpus will used as a foundation to establish a future standard naming and metadata convention for sleep variables in large cohort studies.

Ultimately, we will include sleep as an ontological category in the C-Surv model, a structured taxonomy for harmonizing cohort data, to provide a standardized framework to identify variables of interest, with the ultimate goal to harmonise measures of sleep across different cohort studies. expanding its utility as a universal epidemiological and biomedical ontology ^19^.

## Data Availability

All data and code used will be made available online after peer-reviewed publication.

## Acknowledgements

This work was conducted using the MRC Dementias Platform UK (DPUK). DPUK is a Public Private Partnership funded by the Medical Research Council (MR/L023784/1 and MR/009076/1). For further information on this resource visit www.dementiasplatform.uk.

UB was supported by the Dementia Research Institute through DRI CRT to DJD.

## Supplementary Information

**Table S1:**
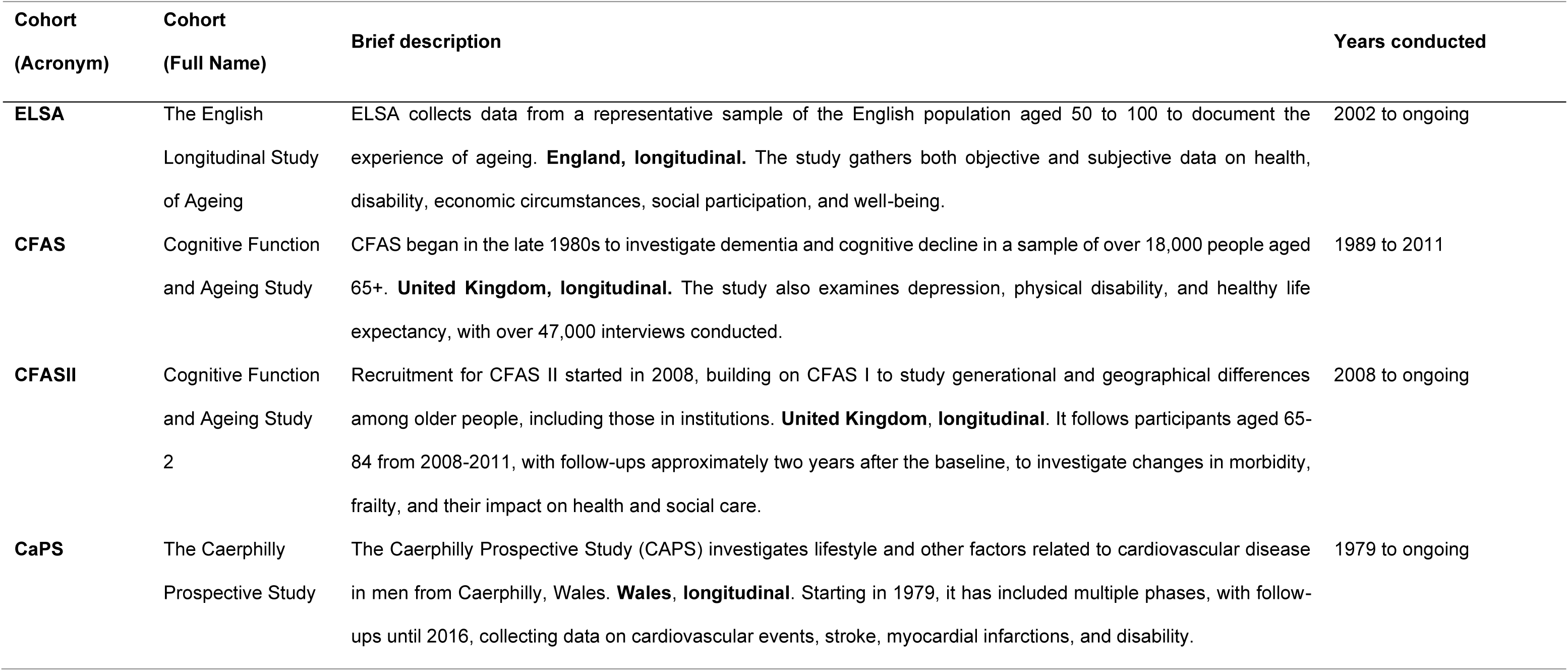
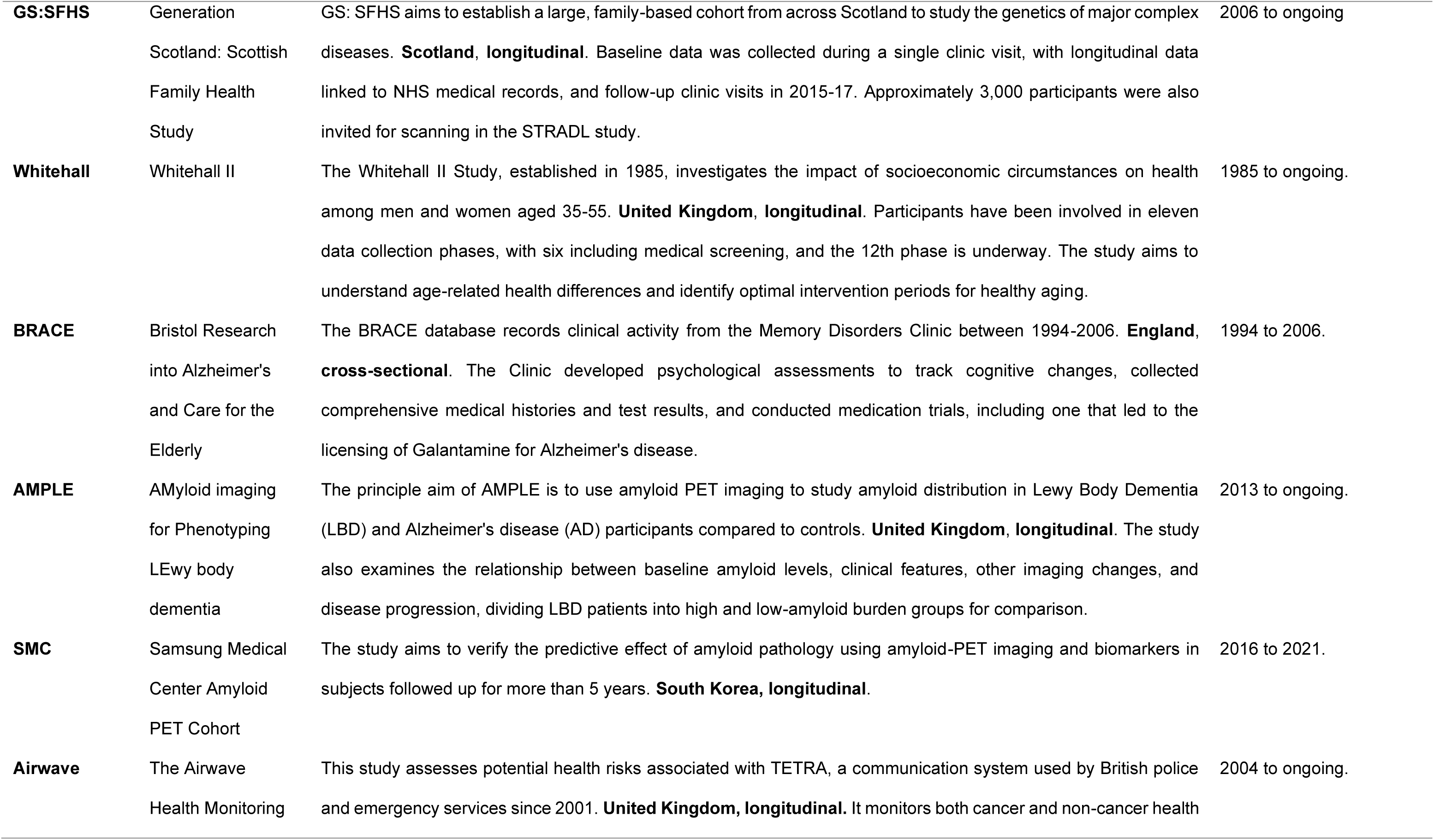
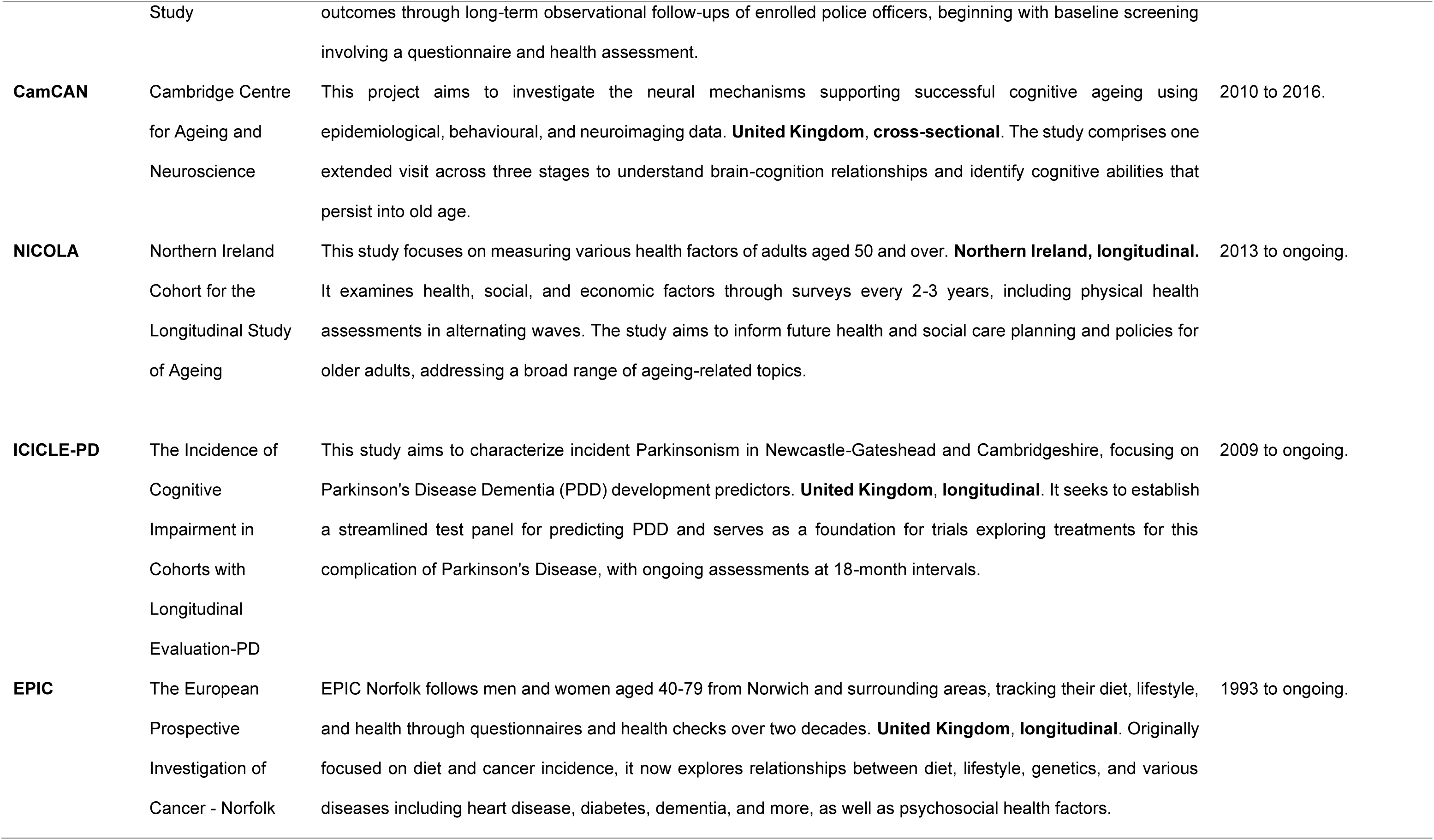
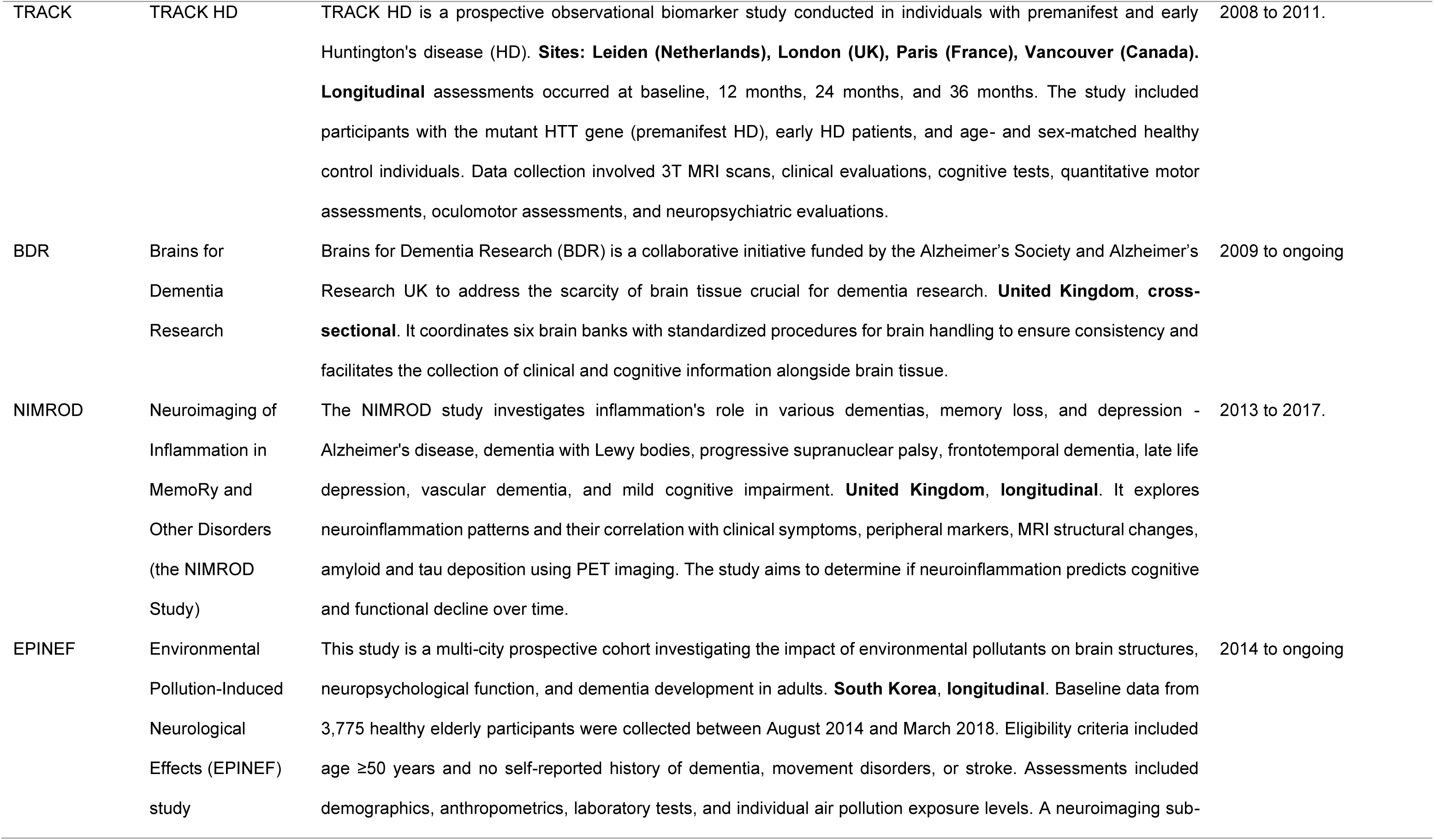
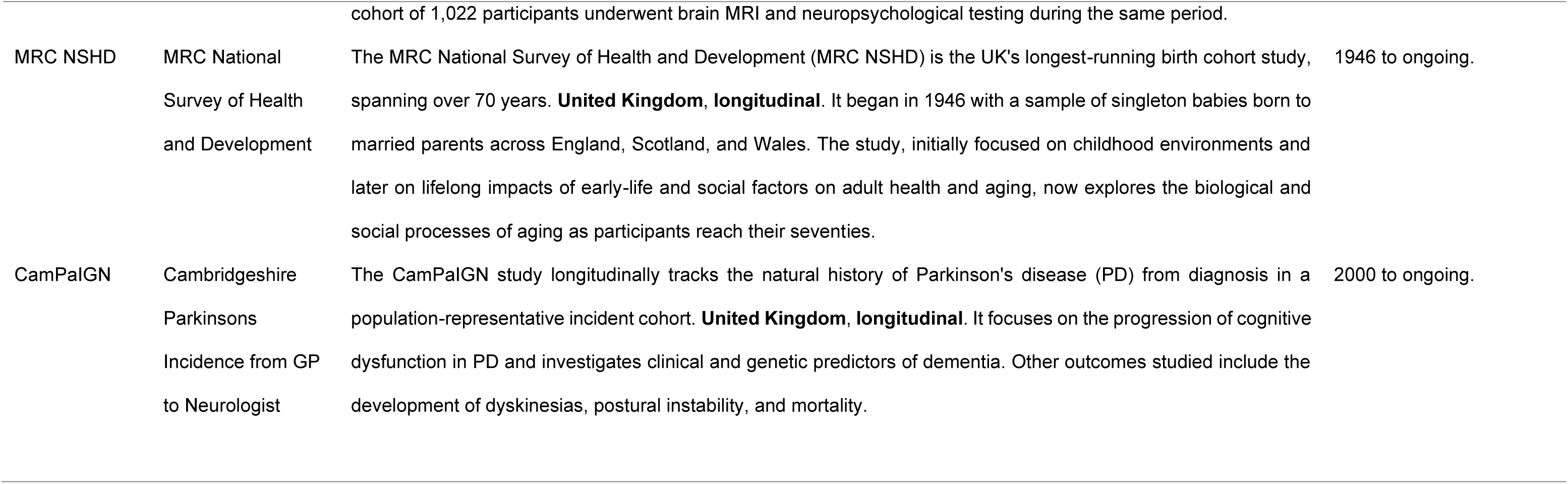
Cohort Information.

| Cohort<br>(Acronym) | Cohort<br>(Full Name) | Brief description | Years conducted |
| --- | --- | --- | --- |
| ELSA | The English<br>Longitudinal Study<br>of Ageing | ELSA collects data from a representative sample of the English population aged 50 to 100 to document the experience of ageing. **England, longitudinal.** The study gathers both objective and subjective data on health, disability, economic circumstances, social participation, and well-being. | 2002 to ongoing |
| CFAS | Cognitive Function<br>and Ageing Study | CFAS began in the late 1980s to investigate dementia and cognitive decline in a sample of over 18,000 people aged 65+. **United Kingdom, longitudinal.** The study also examines depression, physical disability, and healthy life expectancy, with over 47,000 interviews conducted. | 1989 to 2011 |
| CFASII | Cognitive Function<br>and Ageing Study<br>2 | Recruitment for CFAS II started in 2008, building on CFAS I to study generational and geographical differences among older people, including those in institutions. **United Kingdom, longitudinal.** It follows participants aged 65-84 from 2008-2011, with follow-ups approximately two years after the baseline, to investigate changes in morbidity, frailty, and their impact on health and social care. | 2008 to ongoing |
| CaPS | The Caerphilly<br>Prospective Study | The Caerphilly Prospective Study (CAPS) investigates lifestyle and other factors related to cardiovascular disease in men from Caerphilly, Wales. **Wales, longitudinal.** Starting in 1979, it has included multiple phases, with follow-ups until 2016, collecting data on cardiovascular events, stroke, myocardial infarctions, and disability. | 1979 to ongoing |
| **GS:SFHS** | Generation<br>Scotland: Scottish<br>Family Health<br>Study | GS: SFHS aims to establish a large, family-based cohort from across Scotland to study the genetics of major complex diseases. **Scotland, longitudinal**. Baseline data was collected during a single clinic visit, with longitudinal data linked to NHS medical records, and follow-up clinic visits in 2015-17. Approximately 3,000 participants were also invited for scanning in the STRADL study. | 2006 to ongoing |
| **Whitehall** | Whitehall II | The Whitehall II Study, established in 1985, investigates the impact of socioeconomic circumstances on health among men and women aged 35-55. **United Kingdom, longitudinal**. Participants have been involved in eleven data collection phases, with six including medical screening, and the 12th phase is underway. The study aims to understand age-related health differences and identify optimal intervention periods for healthy aging. | 1985 to ongoing. |
| **BRACE** | Bristol Research<br>into Alzheimer's<br>and Care for the<br>Elderly | The BRACE database records clinical activity from the Memory Disorders Clinic between 1994-2006. **England, cross-sectional**. The Clinic developed psychological assessments to track cognitive changes, collected comprehensive medical histories and test results, and conducted medication trials, including one that led to the licensing of Galantamine for Alzheimer's disease. | 1994 to 2006. |
| **AMPLE** | AMyloid imaging<br>for Phenotyping<br>LEwy body<br>dementia | The principle aim of AMPLE is to use amyloid PET imaging to study amyloid distribution in Lewy Body Dementia (LBD) and Alzheimer's disease (AD) participants compared to controls. **United Kingdom, longitudinal**. The study also examines the relationship between baseline amyloid levels, clinical features, other imaging changes, and disease progression, dividing LBD patients into high and low-amyloid burden groups for comparison. | 2013 to ongoing. |
| **SMC** | Samsung Medical<br>Center Amyloid<br>PET Cohort | The study aims to verify the predictive effect of amyloid pathology using amyloid-PET imaging and biomarkers in subjects followed up for more than 5 years. **South Korea, longitudinal**. | 2016 to 2021. |
| **Airwave** | The Airwave<br>Health Monitoring | This study assesses potential health risks associated with TETRA, a communication system used by British police and emergency services since 2001. **United Kingdom, longitudinal**. It monitors both cancer and non-cancer health | 2004 to ongoing. |
|  | Study | outcomes through long-term observational follow-ups of enrolled police officers, beginning with baseline screening involving a questionnaire and health assessment. |  |
| **CamCAN** | Cambridge Centre for Ageing and Neuroscience | This project aims to investigate the neural mechanisms supporting successful cognitive ageing using epidemiological, behavioural, and neuroimaging data. **United Kingdom, cross-sectional.** The study comprises one extended visit across three stages to understand brain-cognition relationships and identify cognitive abilities that persist into old age. | 2010 to 2016. |
| **NICOLA** | Northern Ireland Cohort for the Longitudinal Study of Ageing | This study focuses on measuring various health factors of adults aged 50 and over. **Northern Ireland, longitudinal.** It examines health, social, and economic factors through surveys every 2-3 years, including physical health assessments in alternating waves. The study aims to inform future health and social care planning and policies for older adults, addressing a broad range of ageing-related topics. | 2013 to ongoing. |
| **ICICLE-PD** | The Incidence of Cognitive Impairment in Cohorts with Longitudinal Evaluation-PD | This study aims to characterize incident Parkinsonism in Newcastle-Gateshead and Cambridgeshire, focusing on Parkinson's Disease Dementia (PDD) development predictors. **United Kingdom, longitudinal.** It seeks to establish a streamlined test panel for predicting PDD and serves as a foundation for trials exploring treatments for this complication of Parkinson's Disease, with ongoing assessments at 18-month intervals. | 2009 to ongoing. |
| **EPIC** | The European Prospective Investigation of Cancer - Norfolk | EPIC Norfolk follows men and women aged 40-79 from Norwich and surrounding areas, tracking their diet, lifestyle, and health through questionnaires and health checks over two decades. **United Kingdom, longitudinal.** Originally focused on diet and cancer incidence, it now explores relationships between diet, lifestyle, genetics, and various diseases including heart disease, diabetes, dementia, and more, as well as psychosocial health factors. | 1993 to ongoing. |
| TRACK | TRACK HD | TRACK HD is a prospective observational biomarker study conducted in individuals with premanifest and early Huntington's disease (HD). **Sites: Leiden (Netherlands), London (UK), Paris (France), Vancouver (Canada).** **Longitudinal** assessments occurred at baseline, 12 months, 24 months, and 36 months. The study included participants with the mutant HTT gene (premanifest HD), early HD patients, and age- and sex-matched healthy control individuals. Data collection involved 3T MRI scans, clinical evaluations, cognitive tests, quantitative motor assessments, oculomotor assessments, and neuropsychiatric evaluations. | 2008 to 2011. |
| BDR | Brains for Dementia Research | Brains for Dementia Research (BDR) is a collaborative initiative funded by the Alzheimer's Society and Alzheimer's Research UK to address the scarcity of brain tissue crucial for dementia research. **United Kingdom, cross-sectional.** It coordinates six brain banks with standardized procedures for brain handling to ensure consistency and facilitates the collection of clinical and cognitive information alongside brain tissue. | 2009 to ongoing |
| NIMROD | Neuroimaging of Inflammation in MemoRy and Other Disorders (the NIMROD Study) | The NIMROD study investigates inflammation's role in various dementias, memory loss, and depression - Alzheimer's disease, dementia with Lewy bodies, progressive supranuclear palsy, frontotemporal dementia, late life depression, vascular dementia, and mild cognitive impairment. **United Kingdom, longitudinal.** It explores neuroinflammation patterns and their correlation with clinical symptoms, peripheral markers, MRI structural changes, amyloid and tau deposition using PET imaging. The study aims to determine if neuroinflammation predicts cognitive and functional decline over time. | 2013 to 2017. |
| EPINEF | Environmental Pollution-Induced Neurological Effects (EPINEF) study | This study is a multi-city prospective cohort investigating the impact of environmental pollutants on brain structures, neuropsychological function, and dementia development in adults. **South Korea, longitudinal.** Baseline data from 3,775 healthy elderly participants were collected between August 2014 and March 2018. Eligibility criteria included age ≥50 years and no self-reported history of dementia, movement disorders, or stroke. Assessments included demographics, anthropometrics, laboratory tests, and individual air pollution exposure levels. A neuroimaging sub- cohort of 1,022 participants underwent brain MRI and neuropsychological testing during the same period. | 2014 to ongoing |
| MRC NSHD | MRC National<br>Survey of Health<br>and Development | The MRC National Survey of Health and Development (MRC NSHD) is the UK's longest-running birth cohort study, spanning over 70 years. **United Kingdom, longitudinal.** It began in 1946 with a sample of singleton babies born to married parents across England, Scotland, and Wales. The study, initially focused on childhood environments and later on lifelong impacts of early-life and social factors on adult health and aging, now explores the biological and social processes of aging as participants reach their seventies. | 1946 to ongoing. |
| CamPaIGN | Cambridgeshire<br>Parkinsons<br>Incidence from GP<br>to Neurologist | The CamPaIGN study longitudinally tracks the natural history of Parkinson's disease (PD) from diagnosis in a population-representative incident cohort. **United Kingdom, longitudinal.** It focuses on the progression of cognitive dysfunction in PD and investigates clinical and genetic predictors of dementia. Other outcomes studied include the development of dyskinesias, postural instability, and mortality. | 2000 to ongoing. |

**Table S2:** Cosine Similarity examples for some of the lowest-and highest scoring cohort definitions when matched to UMLS sleep definitions. Lower scoring matches are characterized by partial keyword alignment and the presence of additional contextual or interviewer-specific information, as shown in these longer, more complex definitions. In contrast, higher-scoring matches typically feature shorter, more direct definitions that align more closely with UMLS terms.

| **Cohort** | **variable** | **variable meta data** | **UMLS definition** | **Cosine**<br>**Similarity** |
| --- | --- | --- | --- | --- |
| CFAS |  | s. has difficulty in sleeping? | Difficulty sleeping | 0.960 |
| CFAS |  | trouble sleeping? | Trouble sleeping in past 4W | 0.957 |
| NICOLA |  | my sleep was restless | Sleep is Restless | 0.954 |
| AMPLE |  | sleep disorders - insomnia | sleep disorder; insomnia type | 0.938 |
| CFASII |  | difficulty in getting to sleep | In the past Y, have you had difficulty getting to sleep | 0.886 |
| CFAS | v120_w8 | rating: wakes early by 2 hrs | Does your child wake up early in the morning and have difficulty going back to sleep | 0.296 |
| CaPS | T0532 C5 Self completion quest part2 | [c5 self completion quest q48] how often do you take a nap during the day or early evening? | When you do doze off during the D and take a nap, do you find this sleep refreshing | 0.286 |
| AIRWAVE | F_184 | following traumatic event has been bothered<br>by upsetting dreams | Have you ever had frequent problems getting<br>to sleep at the beginning of the night which<br>has negatively affected how you function<br>during the following day | 0.262 |
| Whitehall | TSLPWAKT | q2.64d wake as usual but tired (s5) | Wake After Sleep Onset | 0.180 |

**Table S3:** Overlapping Categorization of Sleep-Related variables. This table provides a count of variables assigned to each category, distinguishing between generic categories (broadly defined themes) and specific categories (standardized assessment tools). Categories are listed with the number of variables assigned to each, reflecting the distribution seen in the word cloud. Definitions may belong to multiple categories so counts will overlap.

| Category | Count | Group |
| --- | --- | --- |
| Sleep Difficulty | 148 | Generic |
| Sleep Latency and Waking | 77 | Generic |
| Sleep Duration | 64 | Generic |
| Daytime Sleepiness | 41 | Generic |
| Sleep Disorders | 31 | Generic |
| Mental Health and Sleep | 24 | Generic |
| Sleep Environment | 23 | Generic |
| Sleep Quality | 18 | Generic |
| Dreams | 18 | Generic |
| Sleep Medications and Aids | 14 | Generic |
| Physical Health and Sleep | 5 | Generic |
| Alcohol Related | 3 | Generic |
| Sleep Onset | 6 | Generic |
| GHQ | 17 | Specific |
| CESD | 16 | Specific |
| PSQI | 8 | Specific |
| NPI | 6 | Specific |
| Epworth Sleepiness Scale | 2 | Specific |
| Uncategorized | 33 | Generic |

**Table S4:** Full List of Packages and Software used in R and Python. Abbreviations, version number and brief description included.

| Programming Language | Type | Package | Version | Title | Description |
| --- | --- | --- | --- | --- | --- |
| R | Data Transformation | dplyr | 1.1.4 | A Grammar of Data Manipulation | A grammar of data manipulation providing functions to filter, select, mutate, arrange, and summarize data in a fast and intuitive way. |
| R | Data Transformation | tidyr | 1.3.1 | Tidy Messy Data | Tools to tidy data by reshaping it into a consistent and structured format for analysis. |
| R | Data Transformation | readr | 2.1.5 | Read Rectangular Text Data | Efficient tools for reading and writing data files, such as CSV and TSV, into R. |
| R | Data Transformation | stringr | 1.5.1 | Simple, Consistent Wrappers for Common String Operations | Simplifies working with strings, offering functions for pattern matching, manipulation, and analysis. |
| R | Data Transformation | plyr | 1.8.9 | Tools for Splitting, Applying and Combining Data | A versatile toolkit for splitting data, applying functions, and combining results in R. |
| R | Data Transformation | purrr | 1.0.2 | Functional Programming Tools | Tools for functional programming and working with lists, enabling efficient iteration and mapping. |
| R | General Statistics | stats | 4.4.2 | The R Stats Package | A core R package providing statistical analysis tools, including models, tests, and distributions. |
| R | Machine Learning | kernlab | 0.9.33 | Kernel-Based Machine Learning Lab | A comprehensive machine learning package with tools for kernel-based methods like SVM and clustering. |
| R | Natural Language Processing | text | 1.3.0 | Analyses of Text using Transformers Models from HuggingFace, x000D_ Natural Language Processing and Machine Learning | Advanced tools for text mining and natural language processing tasks. |
| R | Natural Language Processing | quanteda | 4.1.0 | Quantitative Analysis of Textual Data | Provides powerful tools for quantitative text analysis and natural language processing. |
| R | Natural Language Processing | udpipe | 0.8.11 | Tokenization, Parts of Speech Tagging, Lemmatization and Dependency Parsing with the 'UDPipe' 'NLP' Toolkit | A natural language processing package that enables part-of-speech tagging, dependency parsing, and lemmatization. |
| R | Natural Language Processing | textrank | 0.3.1 | Summarize Text by Ranking Sentences and Finding Keywords | Implements algorithms like TextRank for keyword and sentence extraction in text data. |
| R | Unsupervised Clustering | Rtsne | 0.17 | T-Distributed Stochastic Neighbor Embedding using a Barnes-Hut Implementation | Implements t-Distributed Stochastic Neighbor Embedding for high-dimensional data visualization. |
| R | Unsupervised Clustering | umap | 0.2.10.0 | Uniform Manifold Approximation and Projection | Implements Uniform Manifold Approximation and Projection for dimensionality reduction. |
| R | Unsupervised Clustering | dbscan | 1.2.0 | Density-Based Spatial Clustering of Applications with Noise_x000D_<br>(DBSCAN) and Related<br>Algorithms | A package for density-based clustering and noise-detection algorithms. |
| R | Unsupervised<br>Clustering | proxy | 0.4.27 | Distance and Similarity<br>Measures | Provides efficient methods for computing distances and<br>similarity metrics between objects. |
| R | Unsupervised<br>Clustering | dendexten<br>d | 1.19.0 | Extending 'dendrogram'<br>Functionality in R | Offers tools for extending and customizing<br>dendrograms in hierarchical clustering. |
| R | Unsupervised<br>Clustering | mclust | 6.1.1 | Gaussian Mixture Modelling<br>for Model-Based<br>Clustering_x000D_<br>Classification, and Density<br>Estimation | Implements model-based clustering, density<br>estimation, and classification using Gaussian mixture<br>models. |
| R | Visualisation | igraph | 2.1.1 | Network Analysis and<br>Visualization | A package for network analysis and visualization of<br>graph structures. |
| R | Visualisation | ggraph | 2.2.1 | An Implementation of Grammar of Graphics for Graphs and Networks | Extends ggplot2 for visualizing networks and graphs in a customizable way. |
| R | Visualisation | ggplot2 | 3.5.1 | Create Elegant Data Visualisations Using the Grammar of Graphics | A widely-used package for creating elegant and flexible data visualizations using the grammar of graphics. |
| R | Visualisation | scales | 1.3.0 | Scale Functions for Visualization | Provides functions for mapping and scaling data in ggplot2, including custom axes and legends. |
| R | Visualisation | ggbreak | 0.1.2 | Set Axis Break for 'ggplot2' | Extends ggplot2 with tools to break axes to handle data with extreme ranges. |
| R | Visualisation | treemapify | 2.5.6 | Draw Treemaps in 'ggplot2' | Creates treemaps using ggplot2 for hierarchical data visualization. |
| R | Visualisation | patchwork | 1.3.0 | The Composer of Plots | Simplifies combining and aligning multiple ggplot2 plots into cohesive layouts. |
| R | Visualisation | wordcloud | 2.6 | Word Clouds | Generates visually appealing word clouds from textual data. |
| R | Visualisation | ggrepel | 0.9.6 | Automatically Position Non-Overlapping Text Labels with_x000D_'ggplot2' | Enhances ggplot2 by adding smartly-placed text and label repulsion for clarity. |
| R | Visualisation | factoextra | 1.0.7 | Extract and Visualize the Results of Multivariate Data Analyses | Simplifies the visualization and interpretation of multivariate analysis and clustering results. |
| R | Visualisation | ggwordcloud | 0.6.2 | A Word Cloud Geom for 'ggplot2' | Integrates word cloud generation into ggplot2 for more customizable text visualizations. |
| R | Python Extension | reticulate | 1.4.0 | Interface to 'Python' | Interface to allow Python Virtual Environments to be run from within an R script. |
| Python | Data Transformation | pandas | 2.2.2 | Powerful Python Data Analysis Toolkit | A versatile toolkit for splitting data, applying functions, and combining results in Python. |
| Python | General Statistics | numpy | 2.2.1 | Fundamental Package for Scientific Computing | A fundamental package for numerical computing in Python, providing support for large multi-dimensional arrays, matrices, and a collection of high-level mathematical functions to operate on these arrays. |
| Python | Natural Language Processing | transformers | 4.44.0 | State-of-the-art Machine Learning for JAX, PyTorch and TensorFlow | A library for natural language processing (NLP) that provides pre-trained models for tasks like text classification, translation, summarization, and more, using transformer-based architectures such as BERT, GPT, and T5. |
| Python | Machine Learning | torch | 2.4.0 | Torch | A deep learning library for Python that provides tools for tensor computation, automatic differentiation, and GPU acceleration, enabling the creation and training of neural networks. |
| Python | Natural Language Processing | polyfuzz | 0.4.2 | Fuzzy String Matching | A Python library for fuzzy matching, designed to compare and cluster text data, providing various algorithms to match similar strings or identify related entities in textual data. |

